## Supplemental Table 1 for "Validation of a risk-prediction model for pediatric post-discharge mortality after hospital admission in Rwanda"

| **Variable** | **Coefficient** |
| --- | --- |
| **Model 1a** | |
| Intercept | -2.760 |
| Weight for age z-score | -0.344 |
| MUAC | -0.295 |
| Sucking well when breastfeeding, or feeding well if not breastfed | -0.248 |
| SpO_2_ | -0.165 |
| Duration of present illness, 48 hours – 7 days | 0.023 |
| Duration of present illness, 8 days – 1 month | 0.194 |
| Duration of present illness, >1 month | 0.068 |
| Age | 0.068 |
| Fontanelle | 0.134 |
| Age × Weight for age z-score | -0.054 |
| Age × Sucking well when breastfeeding | 0.023 |
| Age × Duration of present illness, 48 hours – 7 days | 0.060 |
| Age × Jaundice | 0.101 |
| **Model 1b** | |
| Intercept | -2.753 |
| Weight for age z-score | -0.361 |
| MUAC | -0.219 |
| Time it took to reach hospital, >1 hour | 0.233 |
| Sucking well when breastfeeding, or feeding well if not breastfed | -0.206 |
| SpO_2_ | -0.149 |
| Duration of present illness, 8 days – 1 month | 0.130 |
| Duration of present illness, >1 month | 0.031 |
| Age | 0.031 |
| Age × Weight for age z-score | -0.064 |
| Age × Duration of present illness, 48 hours – 7 days | 0.005 |
| Age × Jaundice | 0.087 |
| **Model 2a** | |
| Intercept | -3.257 |
| MUAC | -0.381 |
| Haemoglobin | -0.225 |
| Weight for age z-score | -0.187 |
| SpO_2_ | -0.176 |
| How long since last admission, <7 days | 0.072 |
| How long since last admission, 7 days – 1 month | 0.132 |
| How long since last admission, 1 month – 1 year | 0.020 |
| How long since last admission, >1 year | -0.058 |
| Water source, bore hole | 0.002 |
| Water source, municipal water | -0.087 |
| HIV+ | 0.102 |
| Age × How long since last admission, 7 days – 1 month | 0.003 |
| Age × How long since last admission, 1 month – 1 year | 0.067 |
| Age × Water source, bore hole | 0.164 |
| Age × Water source, municipal water | -0.036 |
| **Model 2b** | |
| Intercept | -3.243 |
| MUAC | -0.411 |
| SpO_2_ | -0.186 |
| Weight for age z-score | -0.177 |
| Age | 0.029 |
| How long since last admission, <7 days | 0.083 |
| How long since last admission, 7 days – 1 month | 0.142 |
| How long since last admission, 1 month – 1 year | 0.030 |
| How long since last admission, >1 year | -0.081 |
| Respiratory rate | 0.054 |
| Abnormal BCS | 0.156 |
| Temperature, °C | -0.123 |
| Temperature-squared, °C | -0.116 |
| HIV+ | 0.127 |
| Age × MUAC | 0.016 |
| Age × SpO_2_ | 0.011 |
| Age × How long since last admission, <7 days | 0.026 |
| Age × How long since last admission, 7 days – 1 month | 0.023 |
| Age × How long since last admission, 1 month – 1 year | 0.086 |
| Age × How long since last admission >1 year | -0.012 |
| Age × Respiratory rate | 0.106 |
| Age × Abnormal BCS | -0.050 |
| Age × HIV+ | -0.021 |
| **Model 2c** | |
| Intercept | -3.241 |
| MUAC | -0.386 |
| Weight for age z-score | -0.180 |
| SpO_2_ | -0.186 |
| HIV+ | 0.111 |
| Age | 0.007 |
| Water source, municipal water | -0.102 |
| How long since last admission, <7 days | 0.074 |
| How long since last admission, 7 days – 1 month | 0.124 |
| How long since last admission, 1 month – 1 year | 0.013 |
| How long since last admission, >1 year | -0.059 |
| Boil/disinfect/filter water | -0.140 |
| Age × Water source, bore hole | 0.157 |
| Age × Water source, municipal water | -0.044 |
| Age × How long since last admission, <7 days | 0.006 |
| Age × How long since last admission, 7 days – 1 month | 0.007 |
| Age × How long since last admission, 1 month – 1 year | 0.077 |
