## Supplementary figures and images for "Validation of a risk-prediction model for pediatric post-discharge mortality after hospital admission in Rwanda"

### Supplemental Figure 1

#### Hazard curves for post−discharge mortality

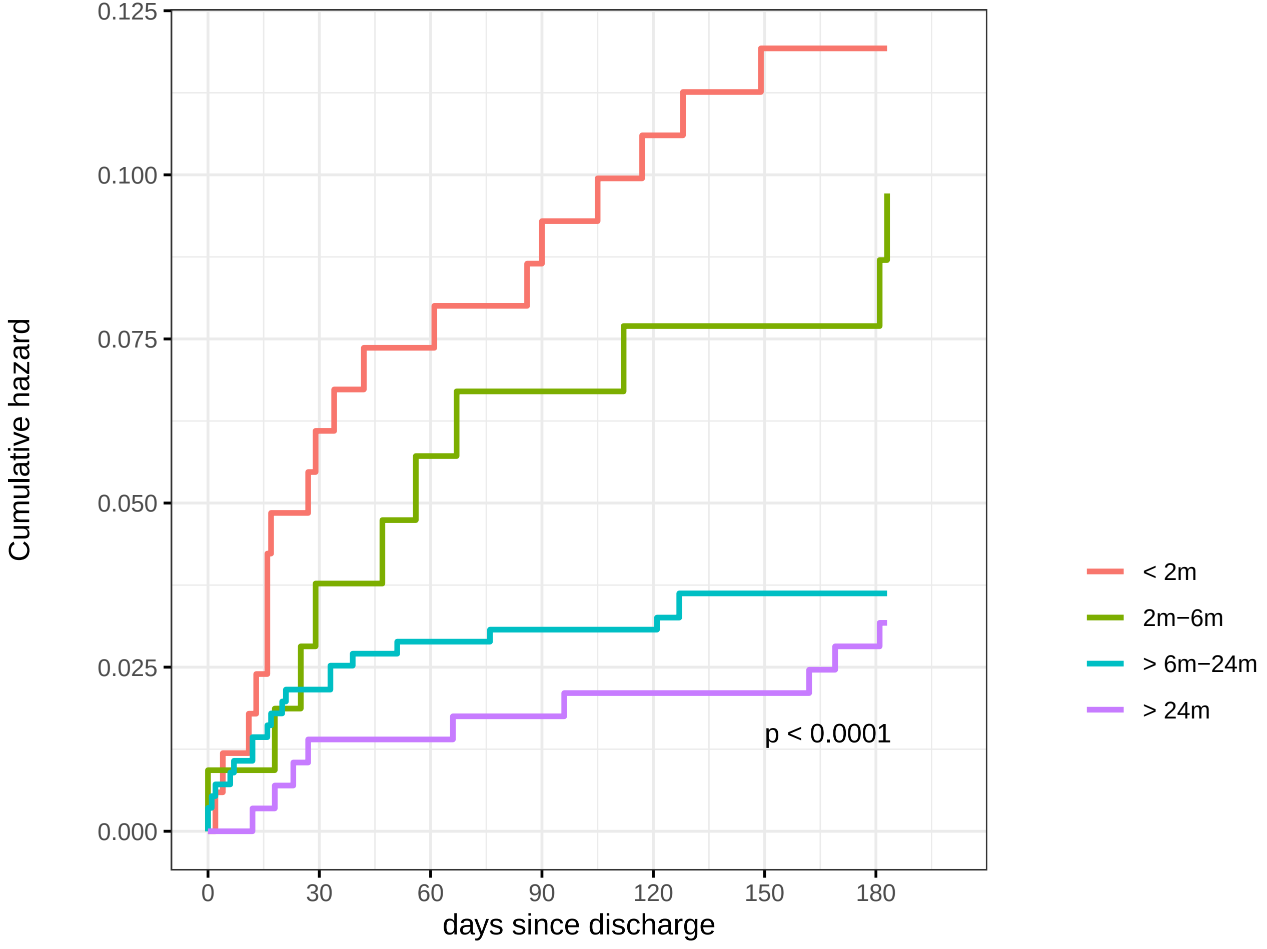


#### Number at risk

| days since discharge | 0 | 30 | 60 | 90 | 120 | 150 | 180 |
|---|---|---|---|---|---|---|---|
| < 2m | 169 | 159 | 157 | 155 | 152 | 150 | 150 |
| 2m−6m | 108 | 104 | 102 | 101 | 100 | 100 | 100 |
| > 6m−24m | 562 | 550 | 546 | 545 | 545 | 542 | 542 |
| > 24m | 288 | 284 | 284 | 283 | 282 | 282 | 280 |

### Supplemental Figure 3

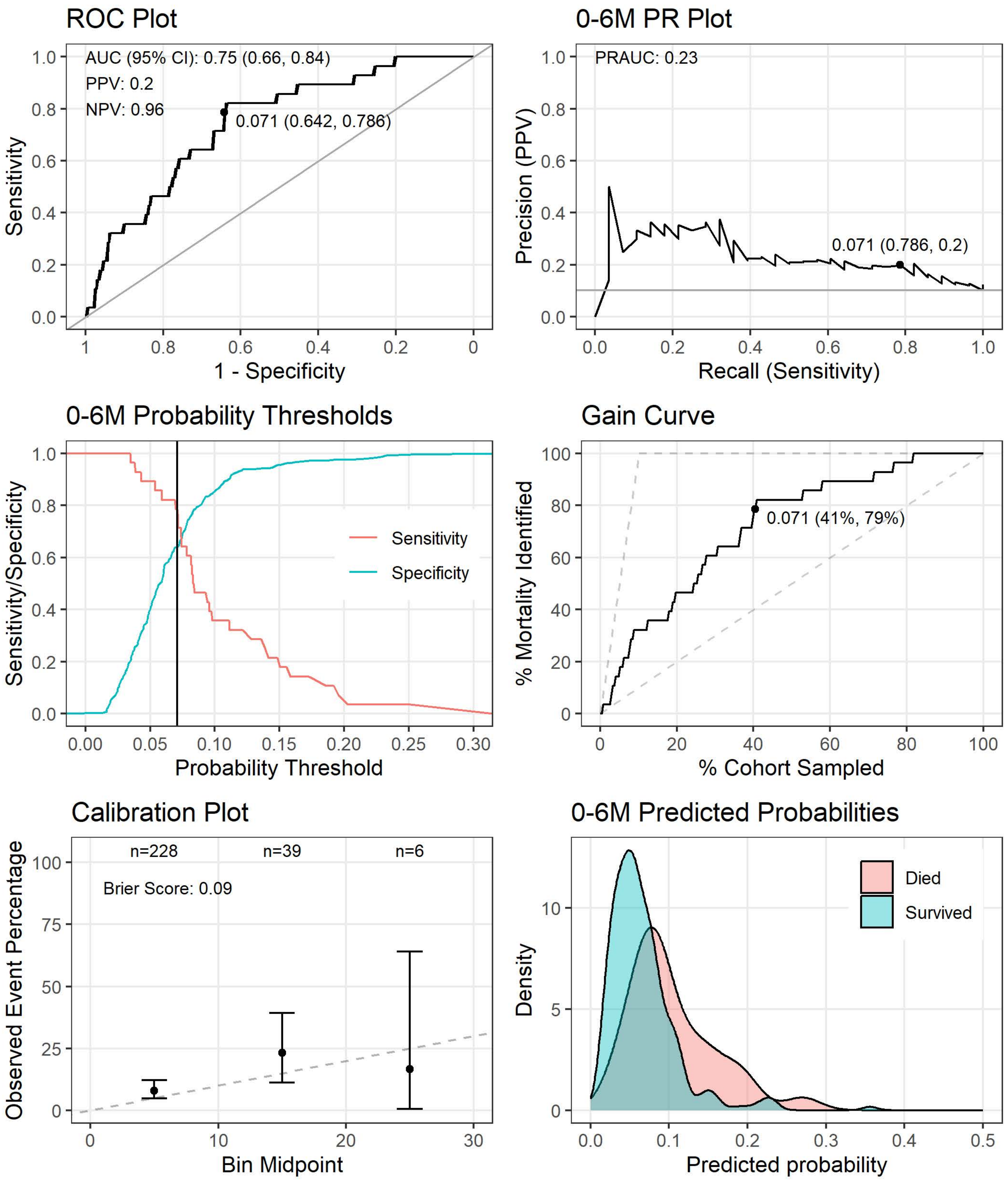

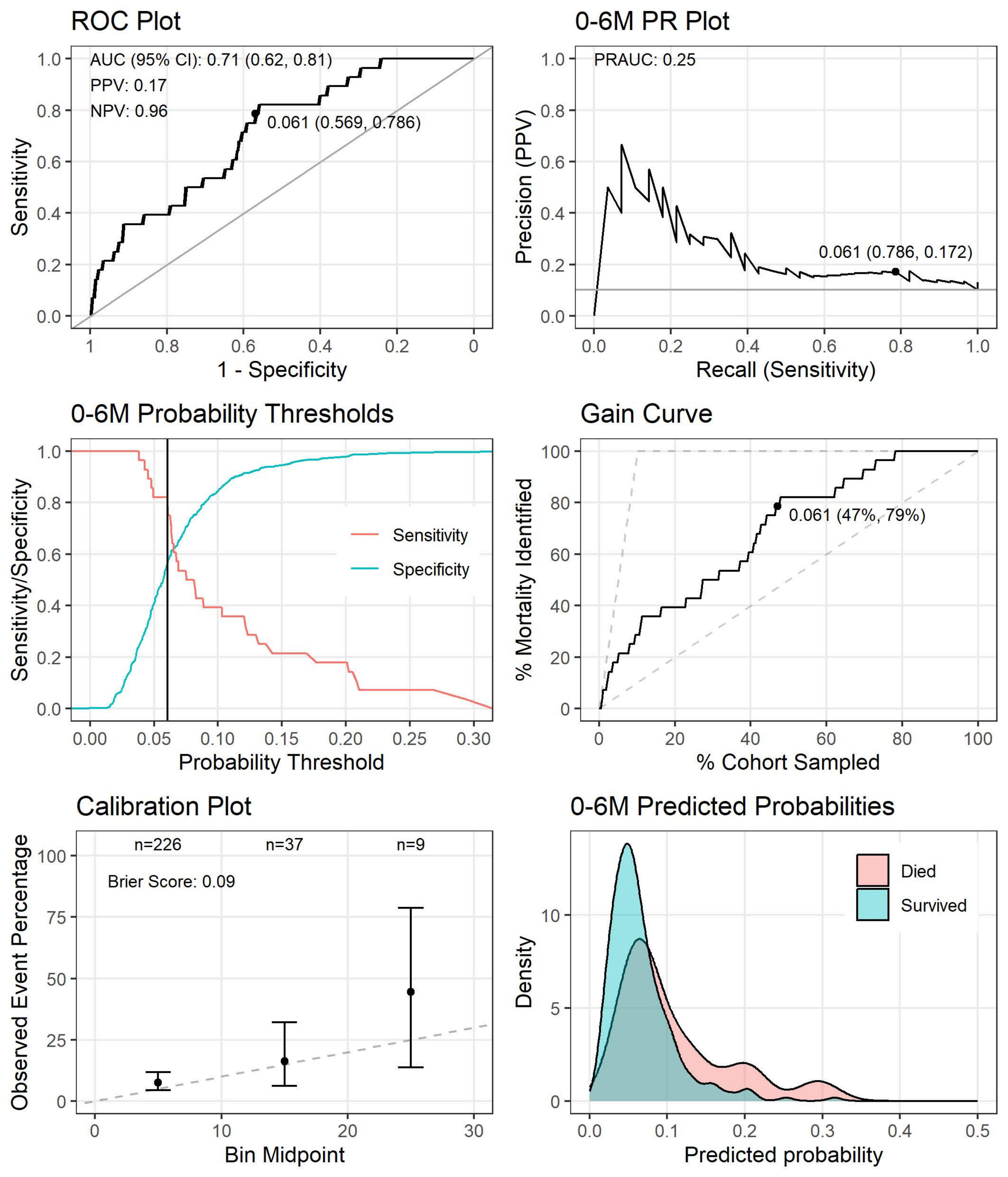

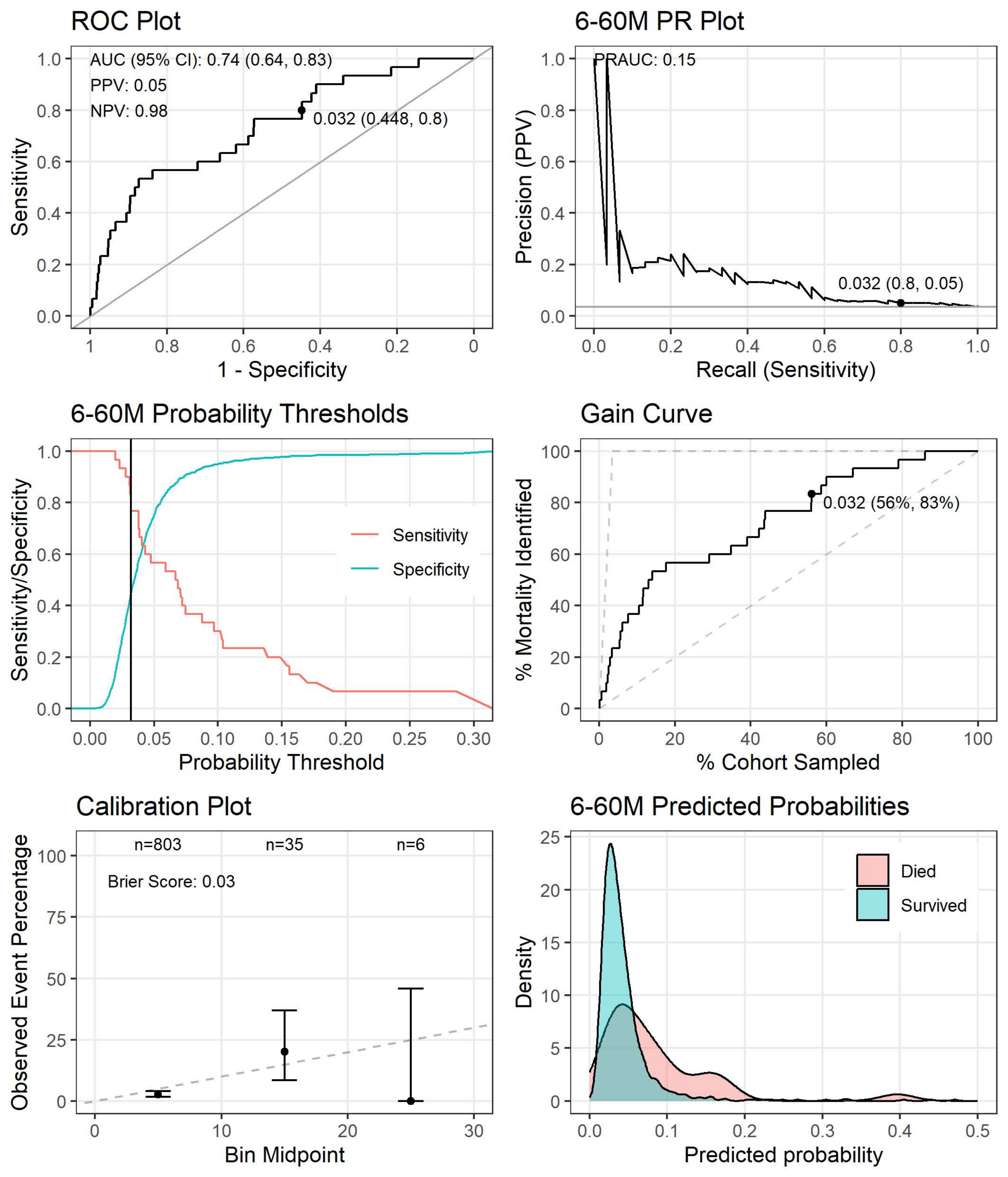

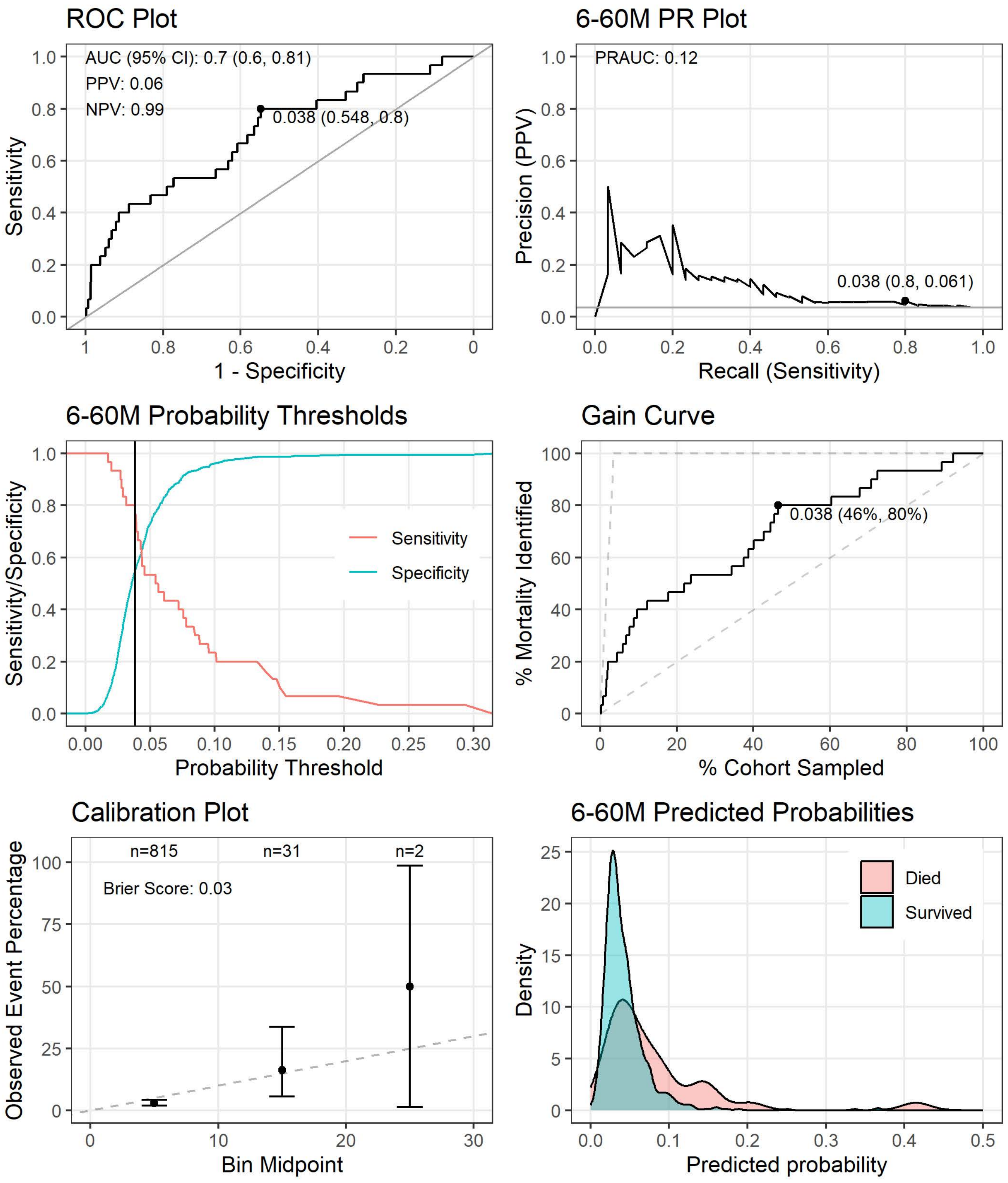

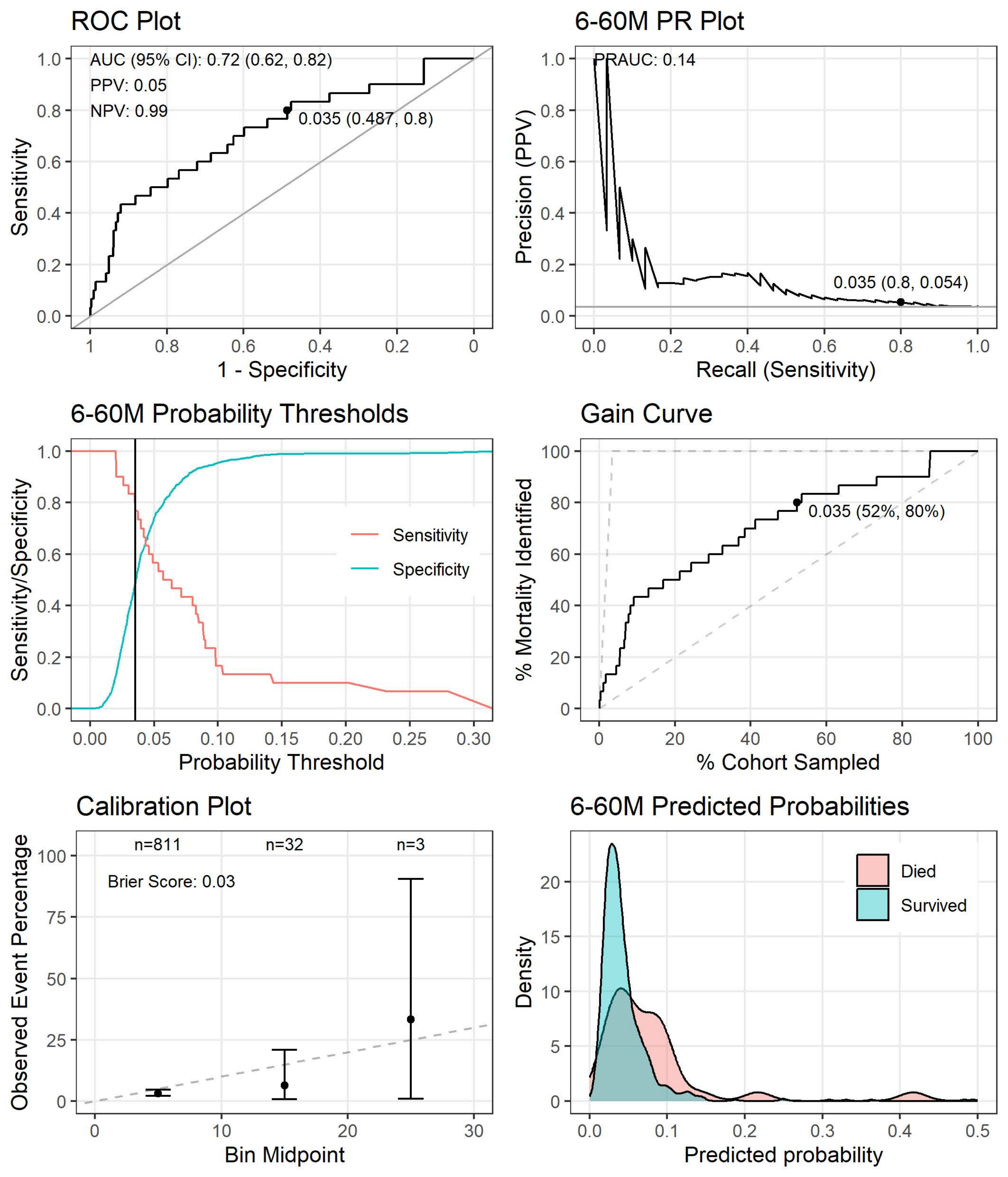
